# Epidemiological Trends and Economic Burden of Genital Warts in Dutch Primary Care

**DOI:** 10.1101/2024.06.03.24307801

**Authors:** C Veijer, J Bes, C Dolk, MJ Postma, LA de Jong

## Abstract

**Background:** This study aims to describe the epidemiological trends and estimate the economic burden of genital warts (GW) in Dutch primary care.

**Methods:** A retrospective, non-interventional, multiyear study (2011-2021) was performed using data from the Nivel Primary Care Database. Changes in incidence by age group, sex, and level of urbanisation of individuals with GW and associated healthcare resource use (general practitioner consultations, prescribed medication, and referrals) were estimated over the 11-year period. Total annual healthcare costs and cost per incident case were estimated via a bottom-up gross costing approach.

**Results:** Between 2011 and 2021, GW incidence increased, which was especially seen in men (from 2.0 to 3.5 per 1 000 inhabitants) and to a lesser extent in women (from 1.9 to 2.1 per 1 000 inhabitants). GW incidence was most common in age group 20-29 years (men: 43.6%; women: 50.7%) and highly urbanised areas. Medication was prescribed in 61.4% of GW cases, and 5.4% of GW patients were referred to secondary care. Total costs in Dutch primary care increased by 108% from EUR 2.3 million in 2011 to EUR 4.9 million in 2021. The cost per incident case also showed an increasing trend from EUR 72 in 2011 to EUR 99 in 2021. Referrals to secondary care resulted in a 14-30% increase of total costs.

**Conclusions:** This study provides novel insights into recent epidemiological trends of GW and its associated costs in Dutch primary care. Incidence especially increased among men and total annual costs of GW in primary care doubled between 2011 and 2021.

## Introduction

Understanding the epidemiological trends and economic burden of a disease is essential when planning preventive measures and efficiently allocating healthcare resources. Especially in the field of infectious diseases, it is useful to evaluate which preventive measures effectively counteract the root cause of disease transmission. One such disease concerns the highly infectious and one of the most common viral sexually transmissible diseases worldwide: genital warts (GW; *condylomata accuminata*).

GW are benign epithelial skin lesions predominantly caused by an infection with non-oncogenic human papillomavirus (HPV) subtypes 6 and 11 [1]. Worldwide incidence rates range between 100 and 200 new cases per 100 000 general adult population, and peak at the age of 15-29 years [2]. The majority of cases are asymptomatic and transient, though recurrence of the self-limiting disease after initial clearance or beyond treatment is common [3]. Treatment options consist of home-based therapy, including self-applied topical treatments podophyllotoxin, imiquimod, and sinecatechins, and clinic-based therapy, including chemical treatments and ablative techniques, such as electrotherapy, cryotherapy, and laser therapy [1].

Apart from pharmaceutical treatment against GW, a number of preventive therapies exists and are globally used to prevent certain HPV-related diseases. Currently, three prophylactic vaccines are licensed in the European Union, all targeting a different range of HPV types: the bivalent vaccine, Cervarix® (targeting HPV types 16/18), the quadrivalent recombinant vaccine, Gardasil® (targeting HPV types 6/11/16/18), and the nonavalent vaccine, Gardasil 9® (targeting HPV types 6/11/16/18/31/33/45/52/58). As of 2010, the Netherlands started routine HPV vaccination for girls only using the bivalent vaccine, which does not provide protection against the HPV types 6/11 causing GW [4].

There is limited data on epidemiology, healthcare resource use (HCRU), and costs of GW in scientific literature. In recent years, a few national studies have described epidemiological trends and have estimated the economic burden of GW [5–11]. Outcome measures differ substantially across these analyses, as well as applied methodologies (e.g. Delphi panel versus physician survey), costing methods (bottom-up micro-costing approach versus claims data), and study setting. Although objectives of the studies are comparable, the methodological variations hamper the generalisability of results to a specific country’s context.

In the Netherlands, the last national study on the economic burden of GW was published two decades ago by van der Meijden *et al.* , who retrospectively analysed treatment patterns, resource use, and costs per patient [12]. The study however did not report any incidence or prevalence estimates of GW in the Netherlands, and population-level costs were not considered. In recent years, new GW incidence rates are roughly estimated at 46 000 episodes per year in the Dutch population, though details on HCRU and costs are lacking [13]. The current study aims to describe the epidemiological trends of patients with GW in terms of age, sex, and level of urbanisation, and to estimate the HCRU and economic burden of GW in the Netherlands.

## Methodology

### Study design and setting

A retrospective, non-interventional, multiyear study was performed, using electronic health record (EHR) data that is routinely recorded by general practitioners (GPs) who participate to the Nivel Primary Care Database (Nivel-PCD), to calculate epidemiological trends and the economic burden of GW between 2011 and 2021 in the Netherlands [14]. This study was approved by the relevant governance bodies of Nivel-PCD (nr. NZR00322.032). According to Dutch legislation, obtaining neither informed consent nor approval by a medical ethics committee is obligatory for this kind of observational study [15].

### Study population

Patients were included if they have been diagnosed at the GP between 2011 and 2021 with *Condylomata accuminata*, based on a registration of the International Classification of Primary Care (ICPC-1) codes Y76 (men) or X91 (women) [16]. Numbers were extrapolated to the total Dutch population based on age, sex, and level of urbanisation. The average number of inhabitants at January 1 of the relevant year was used to calculate each year’s population [17].

### Outcome measures and analysis

Epidemiological (i.e. incidence) and HCRU data (i.e. GP consultations, prescribed drug treatments, and referrals to secondary care) were obtained from the Nivel-PCD, comprising routinely recorded data from EHRs of approximately 500 general practices in the Netherlands (10%) [14]. Routinely recorded data from GPs are an important source of information, as every citizen is registered with a GP and GPs act as gatekeepers in the Dutch healthcare system. The economic burden was calculated by multiplying the HCRU data by unit costs. Unit costs were obtained from various publicly available sources and are described in detail below. Costs were calculated in Microsoft Excel (Microsoft Corporation, Redmond, WA, USA). Data visualisations were produced using Microsoft Power BI (version 2.121.903.0).

To avoid false accuracy, estimates that were based on low numbers were replaced by ‘<100’ for absolute and ‘<0.1’ for relative frequencies before being transferred to Excel. In the analysis, these values were imputed by 50 and 0.05 for absolute and relative frequencies, respectively. This was only applicable to data clustered by age group.

### Epidemiology

Epidemiology measures included incident cases of GW reported at the GP. Incidence is defined as the number of new disease episodes of GW presented at the GP in the relevant year and is reported per 1 000 inhabitants per year. A disease episode was defined as the period between the date of diagnosis and the time of the last encounter plus half of the duration of the contact-free interval. The contact-free interval was defined as the period in which it is likely a patient will visit the GP again if symptoms persist. For GW, the contact-free interval was 16 weeks [18]. Incidence was clustered by sex (men; women), age group (six categories: 0-19; 20-29; 30-39; 40-49; 50-69; ≥70 years), and level of urbanisation (cities: areas with > 1 500 home addresses/km^2^; towns and semi-dense areas: areas with 1 000 to 1 500 home addresses/km^2^; rural areas: areas with < 1 000 home addresses/km^2^).

### Healthcare resource use

GP consultations were clustered by sex, type of consultation, and year. As of January 2019, the categorisation of GP consultation types changed to better cover the increasing trends in consultations by phone and email (SupMat_table1) [19]. Prescribed medication was clustered by sex and year for each type of medication (podophyllotoxin, imiquimod, and sinecatechins). Prescriptions were available as the number of prescriptions per incident case with a prescription and as a percentage of the total number of incident cases. Data on referrals to secondary care were available from 2015 to 2021 and were stratified by type of medical specialty per year.

### Economic burden

The cost analysis used a healthcare payer’s perspective, including direct medical costs only. Costs were calculated using a bottom-up gross costing approach, in which absolute HCRU data were multiplied by unit costs [20]. An overview of unit costs in Euros per year is provided in Supplementary Materials.

Costs of GP consultations were derived from reference prices per consultation type from the Dutch costing manual for economic evaluations in healthcare [21]. The costing manual differentiated GP consultations by two types: standard GP consultation and telephone consultation. The Nivel-PCD data on the number of consultations, though, were differentiated by more than two types (SupMat_table1). Therefore, the cost of telephone consultations were used for email consultations as well, whereas the cost of standard consultation was used for the other types of consultations.

Costs of prescribed medication consist of costs for the pharmacy to purchase the drug, as well as costs incurred as pharmacy service charge (prescription fee) [22]. Pharmacy purchase prices were obtained from the *G-Standaard* [23]. Prices of medication products for GW were selected on the relevant Anatomical Therapeutic Chemical (ATC) classification and included Condylin® (D06BB04 – podophyllotoxin solution for men), Wartec® (D06BB04 – podophyllotoxin cream for women [24]), Aldara® (D06BB10 – imiquimod), and Veregen® (D06BB12 – sinecatechins, introduced in 2012) [25]. The pharmacy service charge was based on the 2020 tariffs for first-time and standard dispensing as reported by the Foundation for Pharmaceutical Statistics (*Stichting Farmaceutische Kengetallen* , *SFK*) [26]. A weighed prescription fee per year was calculated using the average number of prescriptions per patient. Since costs of GP consultations and prescription fees were not available for each year, these were converted to each specific price year (2011-2021) using the consumer price index from Statistics Netherlands [27].

Costs of a referral to a secondary care specialist were based on average selling prices of different hospital products within the relevant diagnosis group of the Dermatology and Venerology specialty (‘*diagnose 21 – SOA* ’) [28]. Prices were obtained from the *‘Diagnose Behandel Combinatie-informatiesysteem’* dataset (a Dutch variant of the Diagnosis-Related Group [DRG] system) of the Dutch healthcare authority (*Nederlandse Zorgautoriteit* , NZa) for secondary care costs [23, 28]. The annual average price per referral visit was calculated by multiplying the share of patients within the diagnosis group by the average selling price per care product that is included in the relevant care product group (‘*Infecties met hoofdzakelijk seksuele overdracht’*).

## Results

### Epidemiology

In the Netherlands, an increase in incidence of GW diagnosed at the GP was observed between 2011 (2.0 per 1 000 inhabitants) and 2021 (2.8 per 1 000 inhabitants). Over the 11-year period, more men than women were newly diagnosed with GW, and the difference increased steadily over time. Male incidence per 1 000 inhabitants increased by 1.5 (2011: 2.0; 2021: 3.5), while female incidence per 1 000 inhabitants increased by 0.3 (2011: 1.9; 2021: 2.2) (Figure 1a). As shown in Figure 1b and SupMat_figure1, incident cases diagnosed at the GP were highest in the 20-29 age group in both sexes (men: 43.6%; women: 50.7%). Incidence across age groups remained stable over the study period. The proportion of men was higher in all age groups except for the youngest age group (0-19 years). Considering incidence of GW by level of urbanisation, incident cases were highest in cities, followed by towns/semi-dense areas and rural areas (11-year averages of 3.3, 2.1, and 1.5 per 1 000 inhabitants, respectively) (Figure 1c). This distribution remained relatively stable over the years.

**Figure 1.**
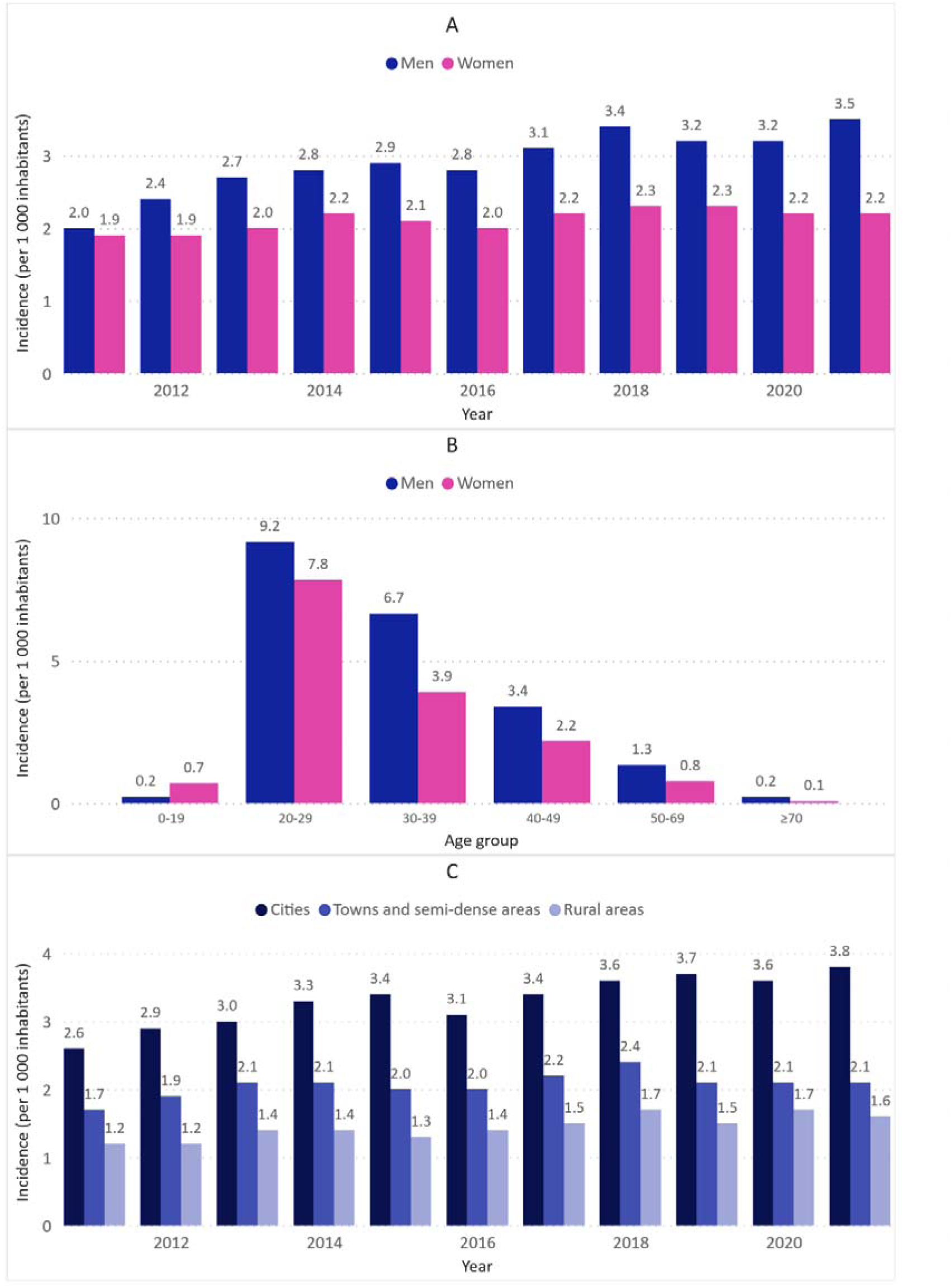
Epidemiology measures of genital warts (GW) based on routinely recorded data from general practitioners (GPs) in the Netherlands between 2011 and 2021. **A) Incidence per 1 000 inhabitants by sex per year; B) Mean incidence per 1 000 inhabitants over the period 2011-2021 by sex and age group** (reported cases for women in the age group ‘≥70 years’ could not be retrieved because of very low numbers and were imputed by 0.05 per 1 000 inhabitants); **C) Incidence per 1 000 inhabitants by level of urbanisation per year**.

### Healthcare resource use

The number of GP consultations per incident case increased from 1.3 in 2011 to 1.6 in 2021 (average: 1.4; men: 1.3; women: 1.5) (Figure 2). On average, 61.4% of GW cases received a prescription for at least one topical treatment (men: 64.0%; women: 58.1%) (Figure 3). Of the total number of prescriptions, podophyllotoxin was the most commonly prescribed type of medication (80.6%), followed by imiquimod (16.1%) and sinecatechins (3.4%) (Figure 4a). The average number of prescriptions per incident case that received a prescription was 1.4 for podophyllotoxin, 1.5 for imiquimod, and 1.5 for sinecatechins (Figure 4b). In the period 2015—2021, on average 5.4% of incident cases were referred to a secondary care specialist. The majority of referrals was made to a dermatologist (75.5%), followed by a specialist at the obstetrics and gynaecology department (17.6%) (SupMat_table3).

**Figure 2.**
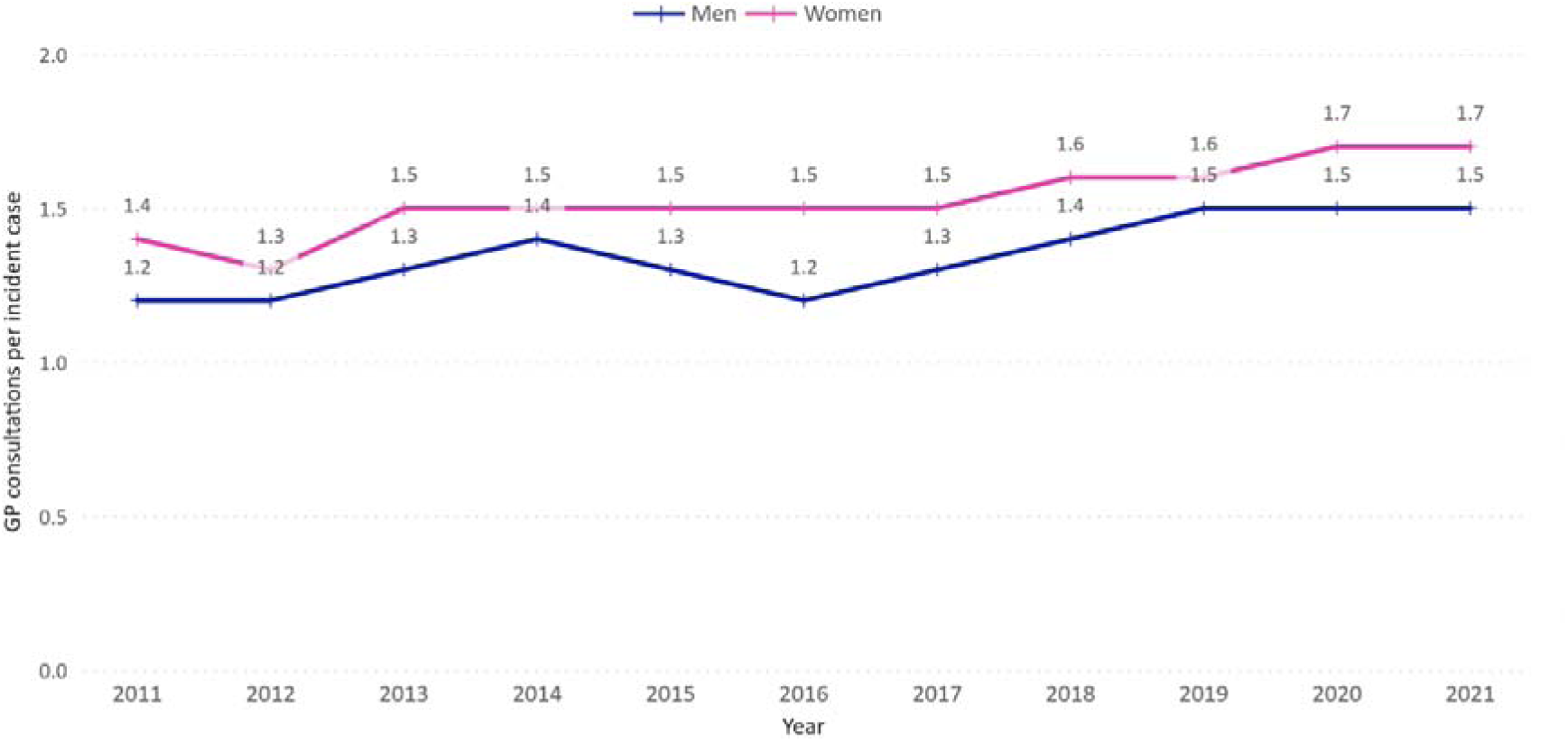
General practitioner (GP) consultations per incident case of genital warts in the Netherlands by sex.

**Figure 3.**
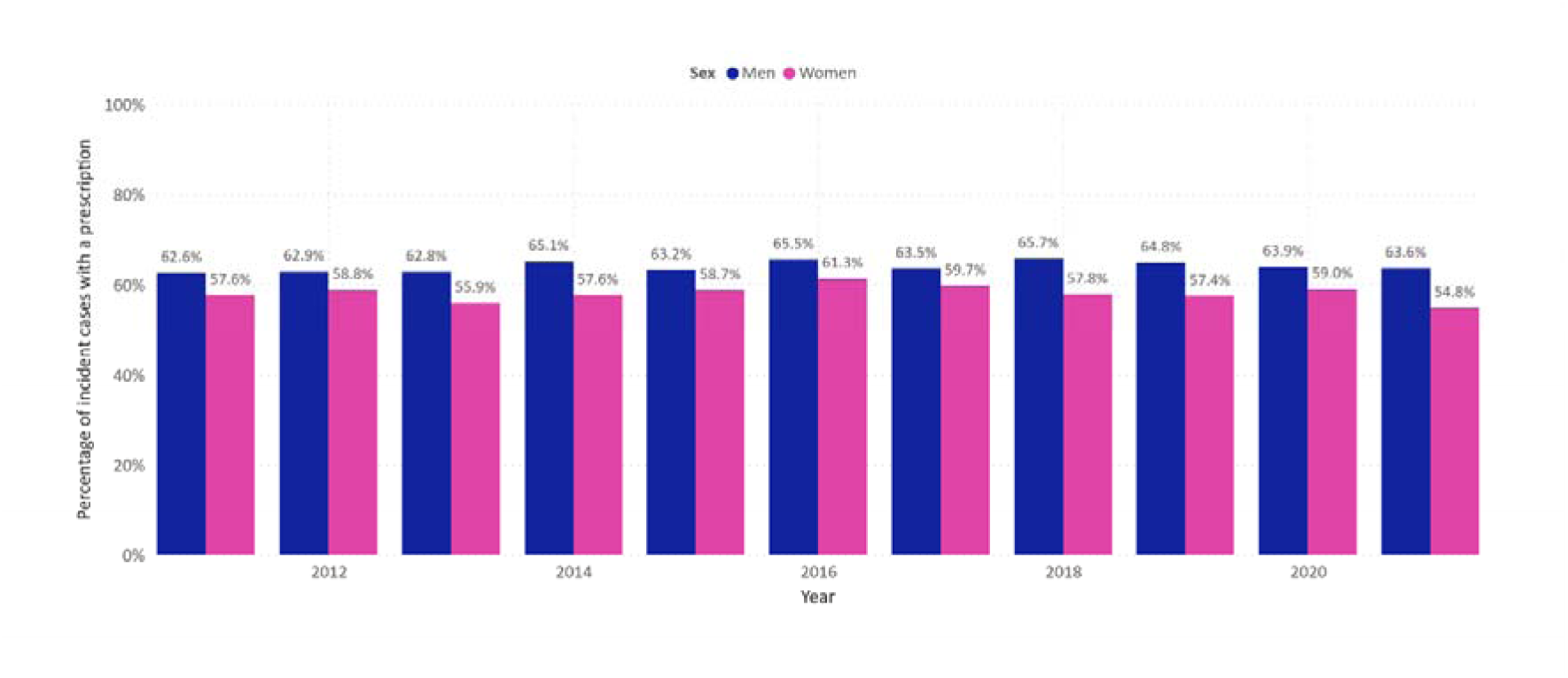
Incident cases of genital warts with a medication prescription as percentage of the total number of incident cases of genital warts in the Netherlands by sex and year.

**Figure 4.**
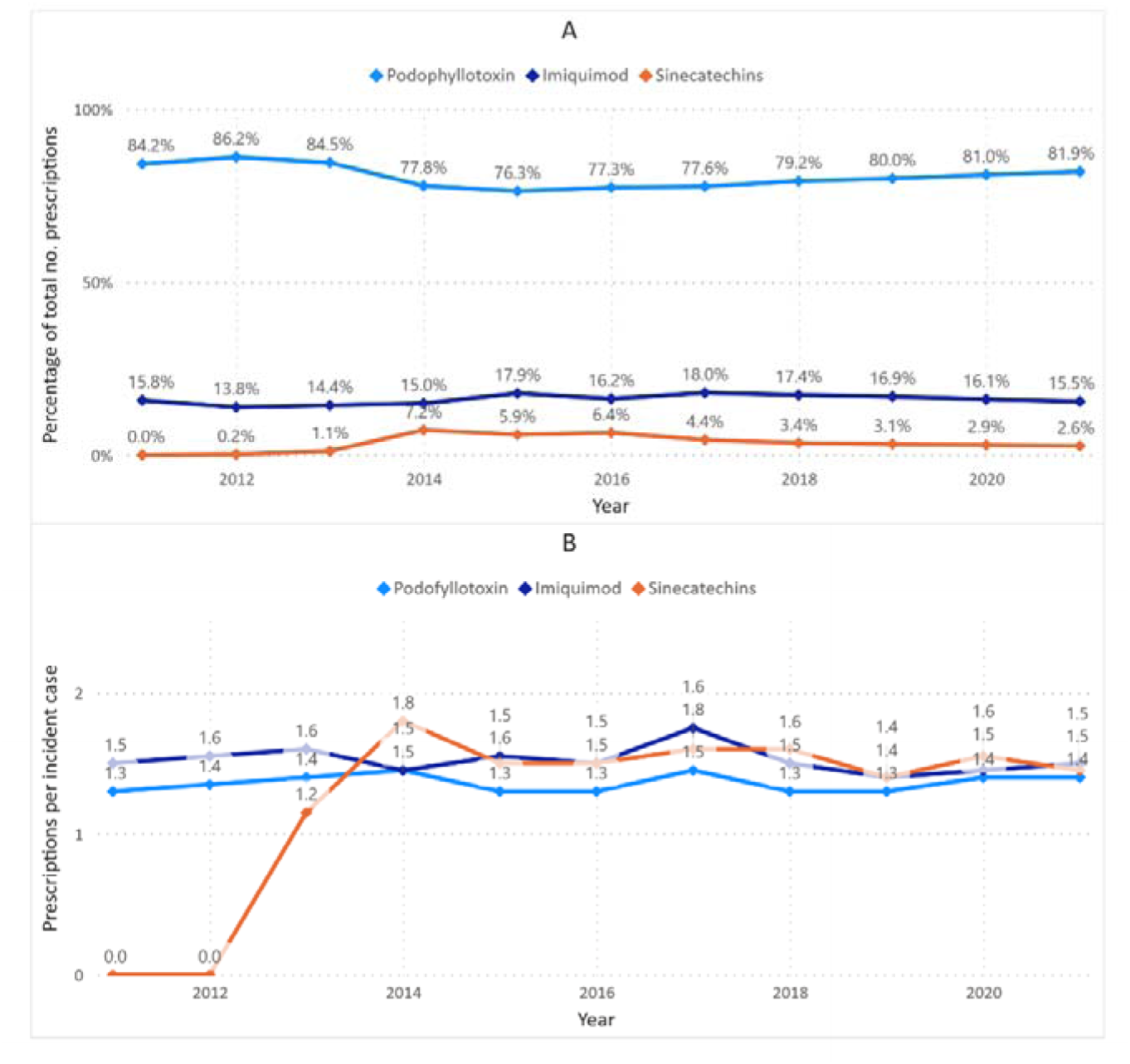
Medication prescriptions for incident cases of genital warts (GW) based on routinely recorded data from general practitioners (GPs) in the Netherlands between 2011 and 2021. A) Prescriptions per type of medication as a percentage of the total number of prescriptions per year for incident cases of GW; B) Prescriptions per incident case of GW with a prescription by type of medication.

### Economic burden

Annual total cost in primary care for GW increased from EUR 2.3 million in 2011 to EUR 4.9 million in 2021. Total costs including costs related to referrals to secondary care increased from EUR 3.9 million in 2015 to EUR 5.8 million in 2021. Costs of GW per year increased by 14 – 30% after adding costs of referrals to secondary care. As of 2015, GP consultations made up the majority of total costs (47.4%), followed by prescribed medication costs (36.6%), and referrals to secondary care (16.0%). Costs per incident case also increased (Table 1). In primary care the cost per case increased from EUR 71.80 in 2011 to EUR 98.76 in 2021. The cost per case including referrals to secondary care increased from EUR 93.29 in 2015 to EUR 117.40 in 2021.

**Table 1.** Direct medical costs of GW per year and costs per incident case of GW per year for only primary care as well as both primary care and referrals to secondary care. Data on referrals to secondary care were available from 2015 to 2021. Abbreviations: GP = general practitioner; GW = genital warts.

| Year | Primary care |  |  |  | Primary care and referrals to secondary care |  |  |
| --- | --- | --- | --- | --- | --- | --- | --- |
|  | GP consultations (€) | Prescribed medication (€) | Total primary care (€) | Costs per GW case in primary care (€) | Referrals to secondary care (€) | Total including secondary care (€) | Costs per GW case including secondary care (€) |
| 2011 | 1,297,062 | 1,052,498 | **2,349,560** | 71.80 | - | - | - |
| 2012 | 1,342,027 | 1,147,382 | **2,489,409** | 69.95 | - | - | - |
| 2013 | 1,613,942 | 1,312,144 | **2,926,086** | 74.79 | - | - | - |
| 2014 | 1,816,489 | 1,732,373 | **3,548,862** | 85.12 | - | - | - |
| 2015 | 1,870,226 | 1,523,172 | **3,393,399** | 81.15 | 507,364 | **3,900,762** | 93.29 |
| 2016 | 1,705,341 | 1,539,553 | **3,244,895** | 78.70 | 670,860 | **3,915,754** | 94.98 |
| 2017 | 2,019,494 | 2,066,432 | **4,085,927** | 90.05 | 821,403 | **4,907,330** | 108.15 |
| 2018 | 2,290,774 | 1,851,250 | **4,142,024** | 84.79 | 1,246,209 | **5,388,233** | 110.29 |
| 2019 | 2,627,105 | 1,754,582 | **4,381,687** | 92.72 | 680,345 | **5,062,033** | 107.11 |
| 2020 | 2,734,133 | 1,897,807 | **4,631,941** | 97.54 | 642,626 | **5,274,566** | 111.08 |
| 2021 | 2,995,797 | 1,901,086 | **4,896,883** | 98.76 | 924,745 | **5,821,628** | 117.40 |

## Discussion

This study is the first analysis of HCRU and associated costs of GW in the Netherlands. Between 2011 and 2021, the incidence of GW in the Netherlands recorded by GPs gradually increased, especially among men. HCRU of GW cases increased accordingly, and total costs increased mainly due to higher incidence and higher medical costs. In general, the results represent a rising trend in economic burden of GW in primary care. Although a minority of new GW cases were referred by the GP to a secondary care specialist (on average 5.4% of patients), the addition of secondary care costs to primary care costs revealed that these are a substantial part of total costs of GW care.

The increase of incident cases was especially present in men. A recently published report by the RIVM based on Nivel-PCD data showed similar trends: the number of episodes of GW in men increased from 2017 through 2021, while the number of episodes for women remained relatively stable [13]. Nonetheless, the reporting rates presented in the RIVM report were extrapolated to the total number of Dutch residents aged 15-64, in contrast to an extrapolation to the whole Dutch population as was done in our analysis. Since the numerators across both studies are different, a valid comparison of the number of episodes cannot be made.

The question can be asked whether the diverging incidence between men and women may be explained by the protective effect of HPV vaccination. Although HPV-6/11 are phylogenetically not closely related to the oncogenic types, there is an ongoing debate about the protective effect of HPV vaccination against GW. A Dutch cohort study estimated vaccine effectiveness six years after vaccination and did not find an effect of the bivalent vaccine against HPV-6/11 infections [29]. This finding was consistent with several other studies, in which no cross-protective effect on genital HPV-6/11 positivity was observed [30–33]. In contrast, a large population-based study in the Netherlands observed a partially protective effect of 23-40% against GW in women fully vaccinated with the bivalent vaccine compared to unvaccinated women [4]. This was in line with two previous studies performed in the United Kingdom, including the results from the largest randomized controlled trial of the bivalent HPV vaccine [34, 35]. However, findings of a large meta-analysis by Drolet et al. including these studies concluded that—although in some younger women the risk on GW seems lower after bivalent vaccination—there were no significant changes in anogenital warts diagnoses between the pre- and post-vaccination periods of the bivalent vaccine [36]. Other recent studies also reported no decrease in HPV-6/11 prevalence following bivalent vaccine introduction [37, 38]. Thus, based on the currently available literature, the diverging incidence between both sexes as observed here cannot conclusively be explained by the protective effect of bivalent vaccination.

Not unexpectedly, incidence of GW according to the level of urbanisation was found to be highest in cities. In general, people living in highly urbanised areas are younger and likely have more interaction than those living outside these areas. A consistently higher incidence of STIs in highly urbanised areas compared to less urbanised areas was also reported in a Nivel report on newly STI-related GP consultations [39].

Our analysis included data from the first two years of the COVID-19 pandemic (2020-2021). Potentially fewer GP consultations due to limited access to healthcare services could have impacted GW related incidence and costs. However, incidence and costs did not decrease during these two years compared to the preceding years in the analysis. In fact, GW incidence increased in 2020 and 2021 for men. As such, no conclusive effect of the COVID-19 pandemic on GW related incidence and costs could be determined.

Medication was prescribed in 61.4% of newly diagnosed GW cases between 2011 and 2021. Prescription rates over the years were consistently higher for men than for women (men: 64.0%; women: 58.1%; at least one prescription per year), which might potentially be explained by the location of the warts. For men, warts are usually situated at the external genital regions, whereas for women, GW more often appear internally, making self-applied treatment less suitable [40].

Total costs were highest in the final year of the analysis, while the largest annual change in costs was observed in 2017. Apart from a rising number in incidence and subsequent GP consultations, drug costs also increased due to price changes. The average pharmacy purchase price of the podophyllotoxin solution increased by 30.8% within one year (2016: EUR 15.38; 2017: EUR 20.13). In 2018, costs of referrals increased by 51.7% compared to 2017, which was mainly due to an enlarged proportion of referrals (SupMat_table3). A direct reason to explain the higher rate of referrals could not be found. Overall, it is arduous to transfer cost outcomes presented here to other jurisdictions, as differences across countries in *inter alia* vaccination strategies, healthcare systems, and clinical pathways are substantial [41].

This study is the first analysis of HCRU and associated costs of GW in the Netherlands, and one of the few estimating the economic burden of GW in the European region [42–44]. The results are based on a representative sample of the Dutch population visiting the GP [14]. The analysis included cost data of the three most relevant resource items in the context of GW. Adding prescription fees to drug costs provided an accurate illustration of the actual expenses made for prescribed treatments.

Nonetheless, the study bears several limitations that should not be neglected. Reported incidence and HCRU and associated costs are surrounded by uncertainties that may have caused an underestimation of the real-world situation. First, data of the Nivel-PCD included reported disease episodes at the GP, leaving GW diagnoses at sexual health centres (SHCs) aside. However, previous research has shown that only around 3% of the GW diagnoses comes from SHCs, indicating that the vast majority of diagnoses is reported at the GP [13]. Second, many cases may be undetected because of shame or an asymptomatic representation of the disease. A recent study in Catalunya (Spain), estimated that approximately one-fifth of GW cases was not registered, especially for women over the age of 30 [45]. Third, costs of secondary care as calculated here are expected to be much higher in actual clinical practice. The number of referrals represent the single fact of a referral being made by a GP to a secondary care specialist, and do not give insight into the number of visits nor the type of treatment given in secondary care. Moreover, risk of wart recurrence after treatment is substantial (20-30%), especially among immunocompromised patients, such as HIV-positive individuals [40]. As such, the actual economic burden of GW in the Netherlands is expected to be even higher than estimated in the current analysis.

On the other hand, several costs may have been overestimated to some extent. The number of prescribed medications are based on registry data of GPs and may have been overestimated as a consequence of non-dispensing of prescribed medication. A Dutch study showed that approximately 10% of first prescriptions in the dermatological drug class (ATC classification) initiated by a GP was not dispensed at the pharmacy [46]. Therefore, at least the sum of prescription fees may be lower than calculated here. Furthermore, not all referred patients may have actually visited a secondary care specialist. One study performed in the Netherlands reported referral compliance rates of 77% and 90% for patients referred for the male and female genital system, respectively [47]. Referral non-compliance might result in fewer costs incurred for secondary care than calculated here.

Besides the volume component in the calculation of secondary care costs, the price component should be interpreted with caution as well. Prices pertained to the care products used in the Dermatology and Venerology specialty only, thereby leaving the care products in the Obstetrics and Gynaecology specialty aside. Also, the share of patients per care product within the diagnosis group included more than just GW patients, since the diagnosis group applies to STIs in general. Moreover, reimbursement claims of health insurers do not accurately reflect the actual expenses made in secondary care [21].

As the outcome of the analysis on the economic burden of GW is surrounded by multiple uncertainties, further similar analyses require less aggregated data. Individual cost data would serve as a valuable input to estimate the cost-effectiveness of HPV-related therapies. Preferably, a micro-costing bottom-up approach should be used, capturing the procedures performed by GPs (to obtain clinic-based as opposed to home-based treatment), indirect cost items (such as travel costs), and HCRU and unit costs in secondary care (in contrast to an average cost per DRG-like payment).

It is beyond the scope and design of the study to correlate health interventions to the epidemiological data as presented here. Nonetheless, from an economic point of view, all costs associated with GW care are opportunity costs and could have been prevented if the disease were to be eliminated. The transmission of the HPV causing GW is influenced by sexual behaviour and smoking [48]. Several strategies exist to reduce the number of GW infections, including education on healthy (sexual) behaviour and vaccination [49]. A declining incidence of GW is pivotal to abate the economic burden on the Dutch healthcare system.

This study provides novel insights into recent epidemiological trends of GW and its associated HCRU and costs in primary care in the Netherlands. Incidence especially increased among men from 2.0 per 1 000 inhabitants in 2011 to 3.5 per 1 000 inhabitants in 2021, and total annual costs of GW in primary care doubled between 2011 and 2021 and was estimated at approximately EUR 5 million in 2021. The results illustrate the need for effective preventive measures and behavioural awareness aimed at the root cause of GW development.

## Data Availability

All data produced in the present work are contained in the manuscript

