## Supplementary_Materials for "Epidemiological Trends and Economic Burden of Genital Warts in Dutch Primary Care"

**Funding:** This study was funded by Merck Sharp & Dohme LLC, a subsidiary of Merck & Co., Inc., Rahway, NJ, USA.

**Supplementary table 1** General practitioner (GP) consultation types recorded until 2018 and as of 2019.

| Vektis code | Description | 2011-2018 | 2019-2021 |
| --- | --- | --- | --- |
| 12000 | Regular consultation shorter than 20 minutes | ● |  |
| 12001 | Regular consultation of 20 minutes or longer | ● | ● |
| 12004 | Regular consultation by telephone* | ● |  |
| 12007 | Regular consultation by email* | ● |  |
| 12010 | Regular consultation shorter than 5 minutes |  | ● |
| 12011 | Regular consultation between 5 and 20 minutes |  | ● |

*: After 2019 categorised under one of the regular consultations, dependent on the length of time. Source: Vektis [1].

**Supplementary table 2** Unit costs per year for GP consultations, prescribed medication, and referrals to secondary care. All costs are presented in Euros (EUR).

|  | **Reference prices GP consultations** (source: Guideline for conducting economic evaluations in healthcare [2]) | | **Average pharmacy purchase prices per SKU**  (source: National drug database [3]) | | | | **Prescription fee**  (source: Foundation for  Pharmaceutical Statistics [4]) | | **Average costs of a referral with diagnosis 21 – SOA*** (source: Dutch health authority [5]) |
| --- | --- | --- | --- | --- | --- | --- | --- | --- | --- |
| **Year** | **Standard consultation** | **Telephone consultation** | **Podophyllotoxin 0.5% solution (men)** | **Podophyllotoxin 0.15% cream (women)** | **Imiquimod**  **5% cream** | **Sinecatechins 10% ointment** | **First-time prescription** | **Regular prescription** |  |
| **2011** | 31.10 | 16.02 | 13.43 | 22.66 | 61.60 |  | 10.34 | 5.62 | 297.51 |
| **2012** | 31.88 | 16.42 | 13.43 | 22.54 | 59.61 | 45.57 | 10.60 | 5.76 | 305.14 |
| **2013** | 32.67 | 16.83 | 13.43 | 23.07 | 56.25 | 79.90 | 10.87 | 5.91 | 240.79 |
| **2014** | 33.00 | 17.00 | 13.43 | 22.57 | 56.09 | 90.29 | 10.98 | 5.96 | 229.37 |
| **2015** | 33.20 | 17.10 | 13.43 | 24.99 | 56.23 | 45.67 | 11.04 | 6.00 | 206.83 |
| **2016** | 33.30 | 17.15 | 15.38 | 26.31 | 55.98 | 41.73 | 11.07 | 6.02 | 201.82 |
| **2017** | 33.76 | 17.39 | 20.13 | 24.85 | 55.67 | 42.52 | 11.23 | 6.10 | 205.25 |
| **2018** | 34.34 | 17.69 | 19.91 | 24.34 | 53.94 | 42.73 | 11.42 | 6.21 | 210.30 |
| **2019** | 35.23 | - | 19.84 | 23.96 | 53.53 | 42.53 | 11.72 | 6.37 | 209.21 |
| **2020** | 35.69 | - | 19.61 | 24.24 | 53.10 | 42.90 | 11.87 | 6.45 | 206.50 |
| **2021** | 36.65 | - | 18.95 | 24.25 | 53.45 | 43.22 | 12.19 | 6.62 | 215.91 |
| Abbreviations: GP = general practitioner; SKU = Stock Keeping Unit. *: average national selling prices of four hospital products (1 or 2 outpatient consultations, >2 outpatient consultations and hospital treatment, surgery, laser therapy) in the relevant diagnosis group were weighed by the number of unique patients registered for each product per year to calculate an average costs of a referral to secondary care for genital warts. | | | | | | | | | |

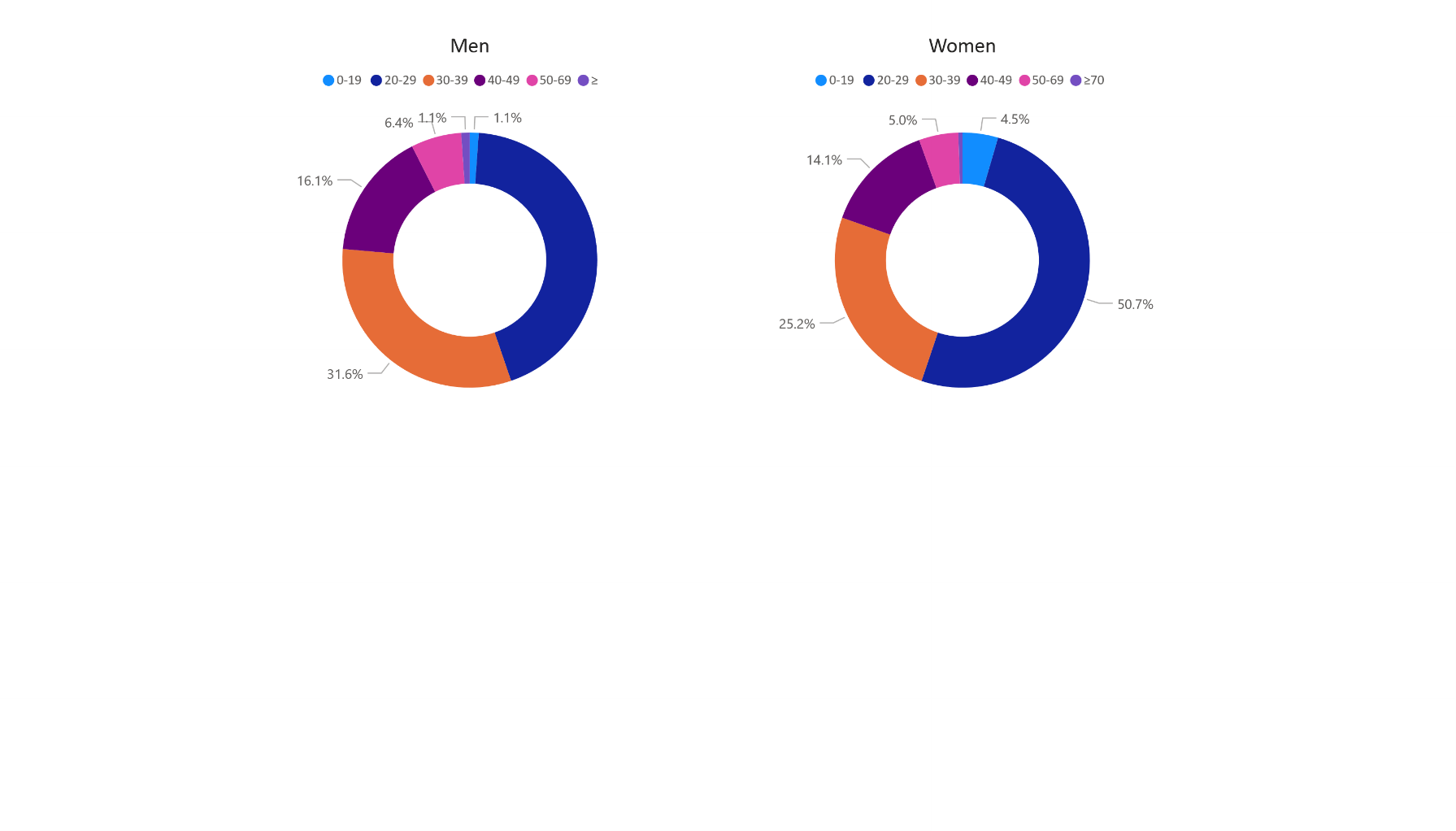

**Supplementary figure 1** Distribution of genital warts incidence per 1 000 inhabitants among six age groups for men and women, based on routinely recorded data from general practitioners (GPs) in the Netherlands between 2011 and 2021. Reported cases for women in the age group ‘≥70 years’ could not be retrieved because of very low numbers and were imputed by 0.05 per 1 000 inhabitants.

**Supplementary table 3** Percentage of incident cases of GW referred by the GP to secondary care and the distribution of referrals among secondary care specialties per year. Abbreviations: GP = general practitioner; GW = genital warts.

| **Year** | **Percentage of incident cases of GW referred to secondary care** | **Medical specialty** | | |
| --- | --- | --- | --- | --- |
|  |  | **Dermatology** | **Obstetrics and Gynaecology** | **Other** |
| 2015 | 4.0% | 65.8% | 30.5% | 3.7% |
| 2016 | 5.9% | 79.5% | 11.5% | 9.1% |
| 2017 | 6.1% | 77.8% | 15.6% | 6.6% |
| 2018 | 8.1% | 75.7% | 20.6% | 3.7% |
| 2019 | 4.4% | 79.2% | 13.9% | 6.9% |
| 2020 | 4.1% | 74.3% | 16.9% | 8.8% |
| 2021 | 5.2% | 75.9% | 14.1% | 10.0% |

2.	Zorginstituut Nederland (ZiNL). Guideline for conducting economic evaluations in healthcare [in Dutch: Richtlijn voor het uitvoeren van economische evaluaties in de gezondheidszorg]. 2016.

3.	Z-Index. Dutch drug database G-Standaard. [Retrieved from: <https://www.z-index.nl/english>.

4.	Stichting Farmaceutische Kengetallen (SFK). Data en feiten 2020 - Het jaar 2019 in cijfers. 2020. <https://www.sfk.nl/publicaties/data-en-feiten/Dataenfeiten2020.pdf>.

5.	Nederlandse Zorgautoriteit. Open data van de Nederlandse Zorgautoriteit. [Retrieved from: <https://www.opendisdata.nl/>.
